# Restarting after COVID-19: A Data-driven Evaluation of Opening Scenarios

**DOI:** 10.1101/2020.05.28.20115980

**Authors:** Ashwin Aravindakshan, Jörn Boehnke, Ehsan Gholami, Ashutosh Nayak

## Abstract

To contain the COVID-19 pandemic, several governments introduced strict Non-Pharmaceutical Interventions (NPI) that restricted movement, public gatherings, national and international travel, and shut down large parts of the economy. Yet, the impact of the enforcement and subsequent loosening of these policies on the spread of COVID-19 is not well understood. Accordingly, we measure the impact of NPI on mitigating disease spread by exploiting the spatio-temporal variations in policy measures across the 16 states of Germany. This quasi-experiment identifies each policy’s effect on reducing disease spread. We adapt the SEIR (Susceptible-Exposed-Infected-Recovered) model for disease propagation to include data on daily confirmed cases, intra- and inter-state movement, and social distancing. By combining the model with measures of policy contributions on mobility reduction, we forecast scenarios for relaxing various types of NPIs. Our model finds that, in Germany, policies that mandated contact restrictions (e.g., movement in public space limited to two persons or people co-living), initial business closures (e.g., restaurant closures), stay-at-home orders (e.g., prohibition of non-essential trips), non-essential services (e.g., florists, museums) and retail outlet closures led to the sharpest drops in movement within and across states. Contact restrictions were the most effective at lowering infection rates, while border closures had only minimal effects at mitigating the spread of the disease, even though cross-border travel might have played a role in seeding the disease in the population. We believe that a deeper understanding of the policy effects on mitigating the spread of COVID-19 allows a more accurate forecast of the disease spread when NPIs are (partially) loosened, and thus also better informs policymakers towards making appropriate decisions.

## Introduction

In response to the COVID-19 pandemic, governments around the world implemented varying degrees of non-pharmaceutical interventions (NPIs) to control the spread of the disease (1,2,3,4). These policies severely restricted movement, public gatherings, national and international travel, and shut down large parts of the economy including schools and non-essential businesses. While the shutdowns helped delay the spread and reduce the severity of the epidemic, they also created tremendous hardships for individuals and businesses (5,6,7). As the spread of COVID-19 decelerated across countries, governments have started relaxing NPIs to help balance the need for economic security and the risk of growing infection numbers (8). Nevertheless, there is limited understanding of the effect that loosening policies might have on the spread of the disease. To determine this effect, we quantify each NPI’s contribution to disease mitigation, permitting the forecasting of disease spread under different policy scenarios. The proposed model, then, will allow policymakers to forecast the impacts of the removal of different types of restrictions.

Initial analysis of the impact of policy restrictions in China suggests that NPIs that significantly affected human mobility (e.g., household quarantine) reduced the spread of the disease (7, 10), even more than restrictions that limited national and international travel (11). Additionally, simulations of NPIs in Wuhan (6) show that maintaining restrictions helped delay the epidemic peak. The results also imply that an early end to such interventions leads to an earlier secondary peak, which can be flattened by relaxing the social mixing at varying rates (6). Nevertheless, to the best of our knowledge, no study quantifies the effects of the types and timings of the implementation and relaxation of government policy interventions in reducing mobility and in turn decreasing the spread of COVID-19. Our estimates allow for projections of the impact of easing individual interventions on disease spread. These predictions act as decision aids for policy makers to judge how lifting certain policies changes social mobility rates and in turn the number of new COVID-19 cases.

Using data from the 16 states of Germany, we explore the effectiveness of different NPIs (Figure 3) in reducing social mobility, and in turn affecting the spread of the disease. Because German states enforced (and relaxed) policies to varying degrees and at different points in time, the variations in implementation allow us to capture the incremental effectiveness of these policies at reducing social mobility amongst the general population.

To determine how policy enforcement impacted mobility and disease spread, we associate the type and timing of the policy intervention to actual social mobility as recorded in the data released by Google (14). Next, using our predictions of social mobility based on the policy interventions, we predict the spread of COVID-19 by modifying the SEIR model presented in (9) to include social distancing and other forms of mobility data (e.g., travel by air, bus, rail, and road). Finally, we project the impact of relaxing a policy on the number of new cases across Germany and compare how differences in start times for policy relaxations alter the cumulative number of expected cases over a six-week time span.

We find that not implementing social distancing in Germany, would have resulted in a 37.8-fold (IQR: 27 to 52-fold) increase in cumulative infected case counts as of May 7, 2020. In other words social distancing reduced case counts by about 97.3% (IQR: 96.3–98.07%). We also find that policies are not equal in their effectiveness at reducing new cases. Contact restrictions were the most effective at lowering infection rates in Germany (51%, IQR: 50.7–51.2), while border closures had only minimal effects at mitigating the spread of the disease (2%, IQR: 0 – 4%), even though they might have played a role in seeding the disease in the population.

## Methods

### Incident Data for COVID-19 cases

Interconnected air, land, and sea transportation networks led to the spreading of COVID-19 from Wuhan, China to the rest of China and eventually to most countries around the world (12, 13). To accurately model the spatial spread of the disease into Germany, we collected three types of daily mobility data: (i) daily air transportation data to capture the movement within and between Germany and 142 other countries; (ii) daily ground transportation data between the nine countries that share borders with Germany; and (iii) daily inter-state ground transportation. The daily COVID-19 case data were obtained from the Johns Hopkins Coronavirus Resource Center (15) and Robert Koch Institute (16) for all countries in our dataset as well as the 16 German states (see Figure 1). Figure 2 shows the cumulative case numbers for all states in Germany and globally.

**Figure 1.**
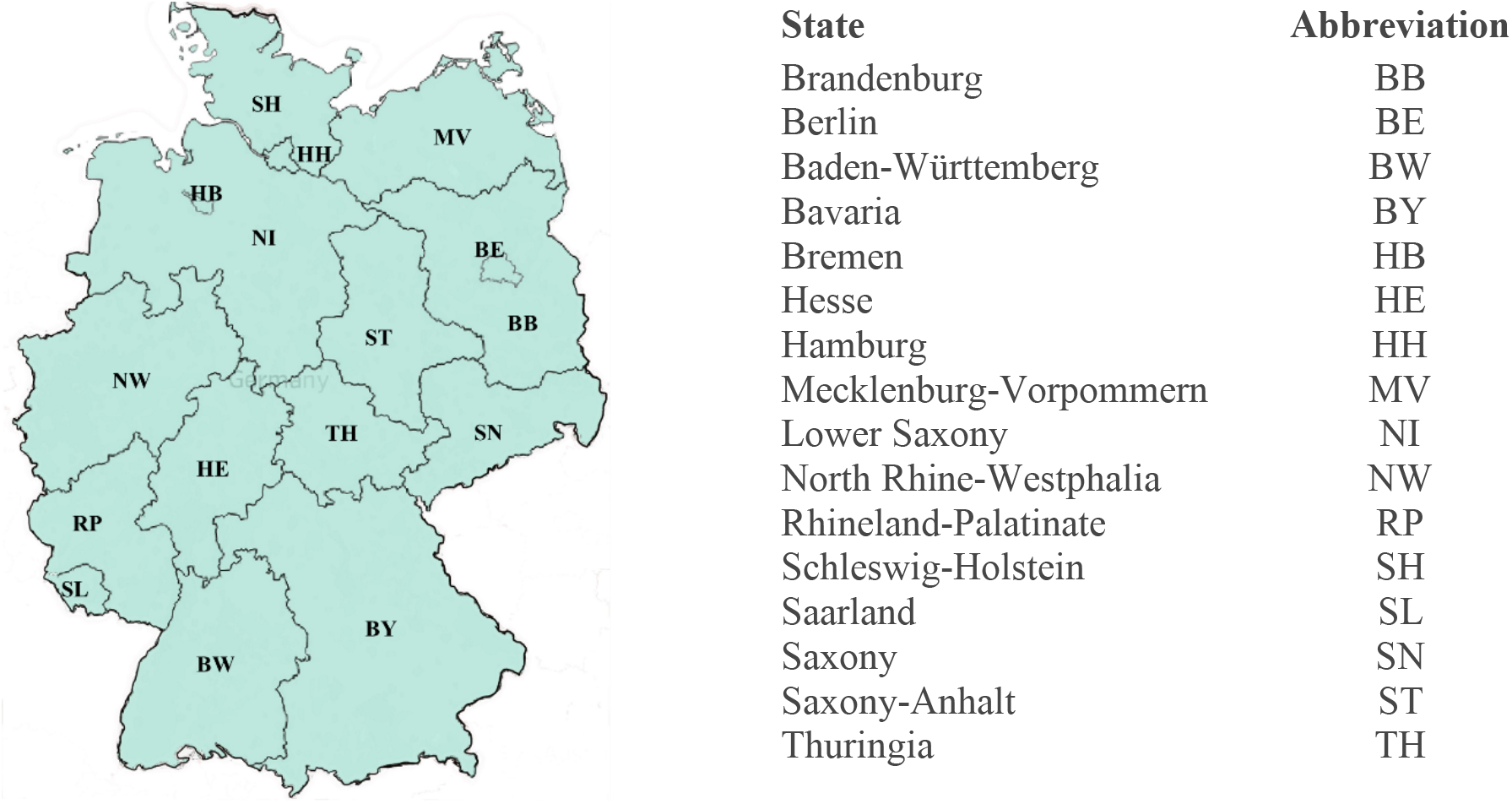
Map of Germany and Abbreviations of the 16 German states.

**Figure 2.**
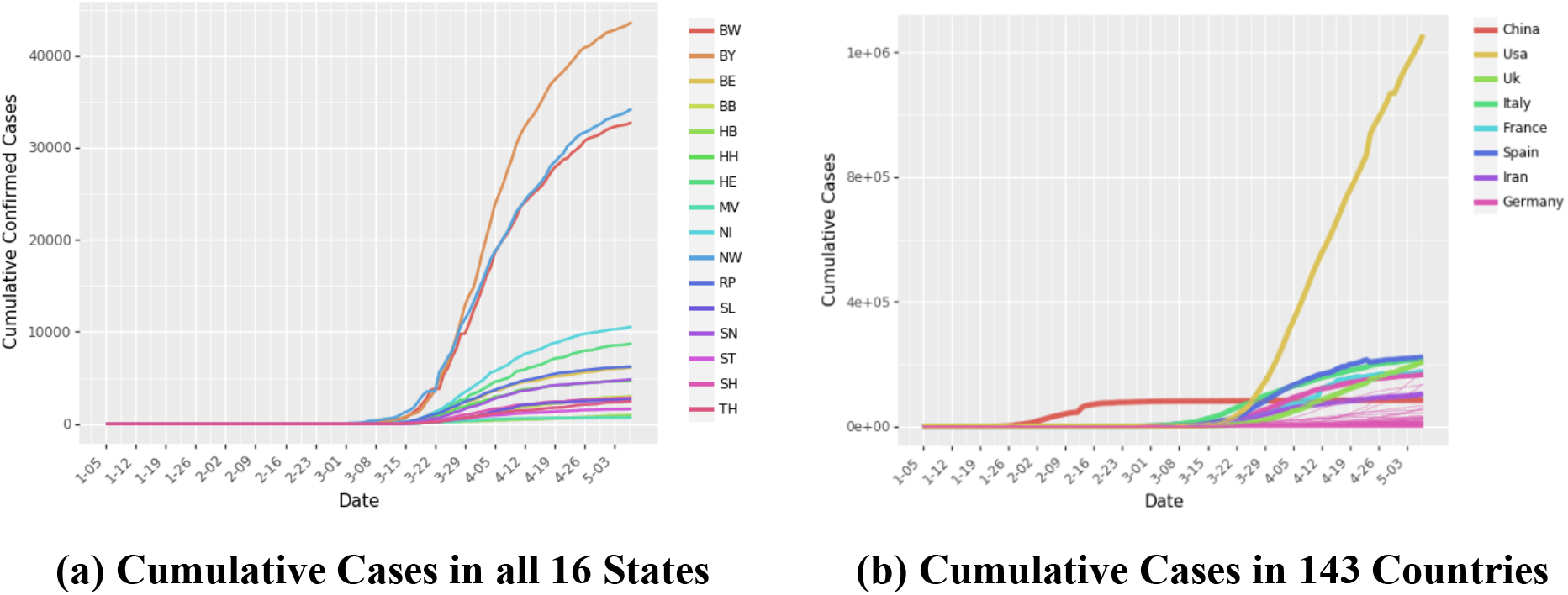
Confirmed positive cases for COVID-19 in Germany and the world. Panel (a) shows the cumulative curves for the 16 states of Germany. Panel (b) shows the cumulative curves from other 142 countries. We highlight the countries with very high number of cases – China, Italy, USA, UK, France, Spain, and Iran.

### Government Policies

To encourage and enforce physical distancing, governments across all 16 states introduced a variety of NPIs at different points in time (Figure 3). Data for these policies were collected from (17, 18). Table S2 in the Supplementary Materials describes the policies implemented across Germany. To understand the impact of each policy in containing the spread of the disease, we analyze (i) what would have happened if individuals did not reduce their mobility, i.e., social distancing norms were never introduced, and (ii) what would happen if a policy *p* is relaxed in the future. While the former can help governments in the early implementation of critical policies in case of future epidemics, the latter can help governments during current and future epidemics to decide which policies to relax first.

**Figure 3.**
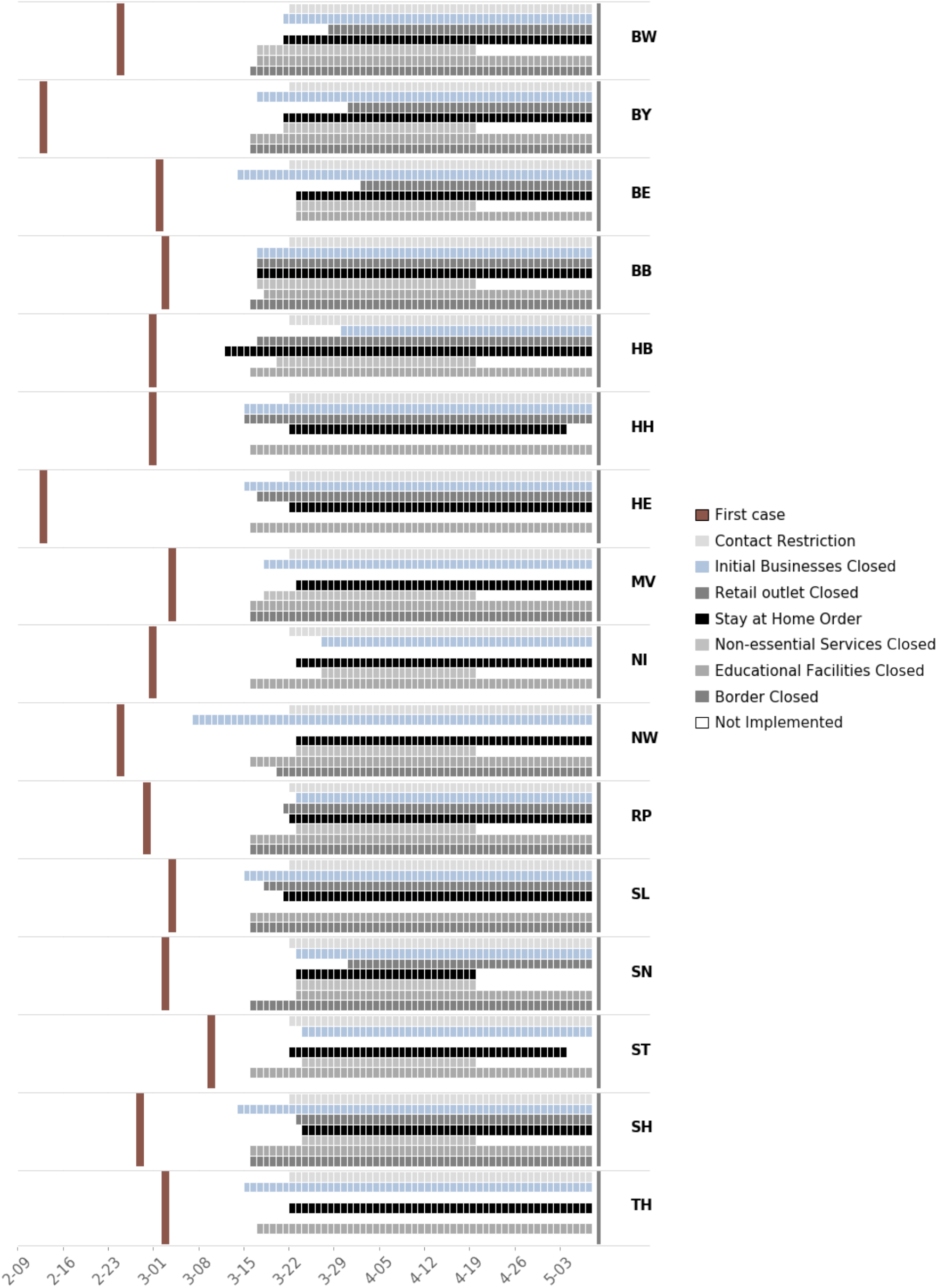
Timeline for implementation of NPIs across states. These policies include border closures (closing international borders), contact restriction (movement in public space is limited to two persons or people co-living), educational institutes closure (e.g. schools and universities), initial business closure (e.g. restaurants), non-essential business closure (e.g. trade shows), stay at home order and retail store closure. Border closure applies to 10 states sharing international borders. We use data from March 1, 2020 to April 30, 2020 in our study. Every policy was not implemented by every state as of April 26, 2020. Also, none of the implemented policies were relaxed until April 20. State governments start relaxing these policies from April 20, 2020.

### Community Mobility

Google’s COVID-19 Community Mobility Reports (14) detail how movement trends change over time as public awareness increases and NPIs are introduced (Figure 4). The report tracks movement trends over time by geography, across different categories of places such as retail and recreation, groceries and pharmacies, parks, transit stations, workplaces, and residential. We consider community mobility trends in retail and recreation as a measure of social distancing. We define social distancing as *sd_i_ = –C_i_/100* where *C_i_* is the community mobility trend in state *i*.

**Figure 4.**
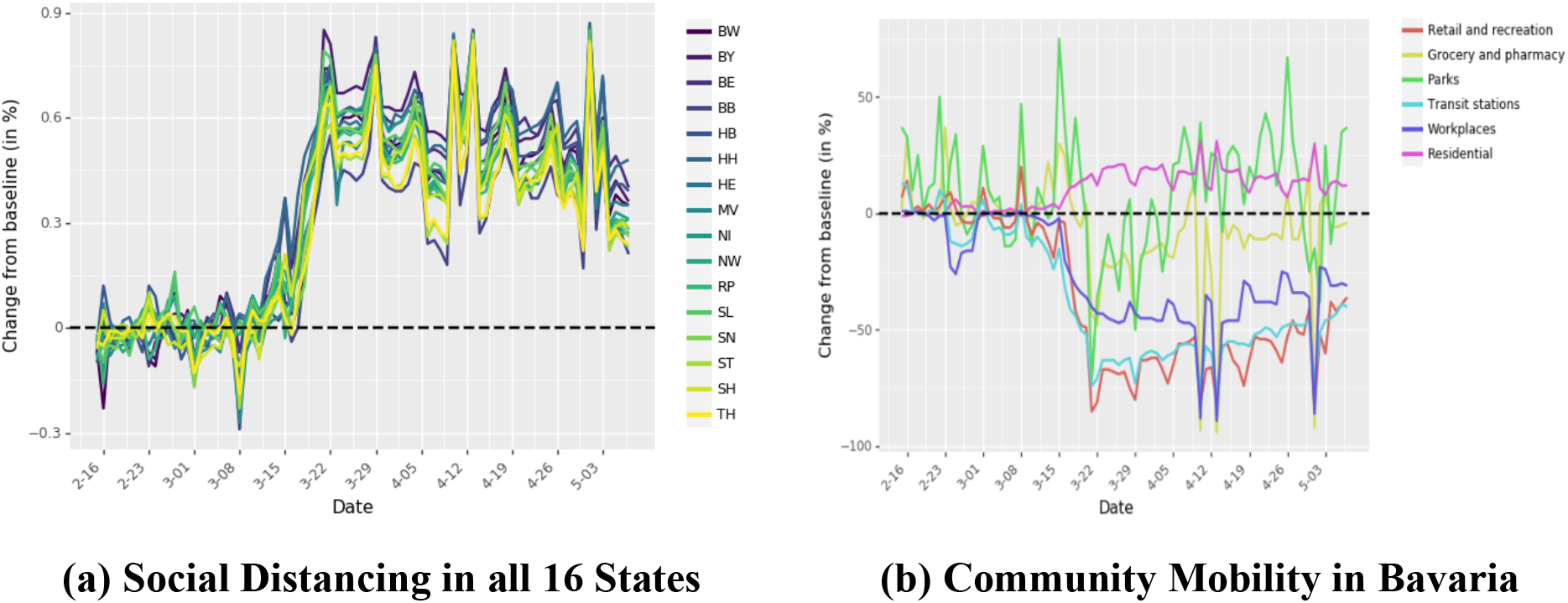
Community mobility over time. Community mobility data charts the difference in foot traffic as compared to baseline from historical data. Panel (a) shows the social distancing over all the 16 states. Panel (b) shows the movement trend across different categories of places for Bavaria. Similar movement trends for all the other states are shown in the Supplementary Material.

While the community mobility data provides information on changes in local movement, it does not provide information on inter-state movement and international travel. Ground transportation accounts for the vast majority of the movement, with cars accounting for 85% of total ground transportation in Germany (19). We collected detailed traffic data from Jan 1, 2013 to Dec 31, 2018 from the German Bundesanstalt für Straßenwesen (Federal Institute for Roadways). The dataset contains the hourly count of the number of vehicles crossing different checkpoints along highways. The institute used sensors to identify the type of vehicle, which we include in our analysis to estimate the number of individuals. We construct a linear regression model to predict hourly traffic for Jan 1, 2020 to April 30, 2020 (details in Supplementary Material). The model includes year, public holidays, day of the week, and state population as control variables. To control for changes in car movement during the period of the study, we adjust the predicted daily traffic using Google’s community mobility data for workplaces (Figure 5).

**Figure 5.**
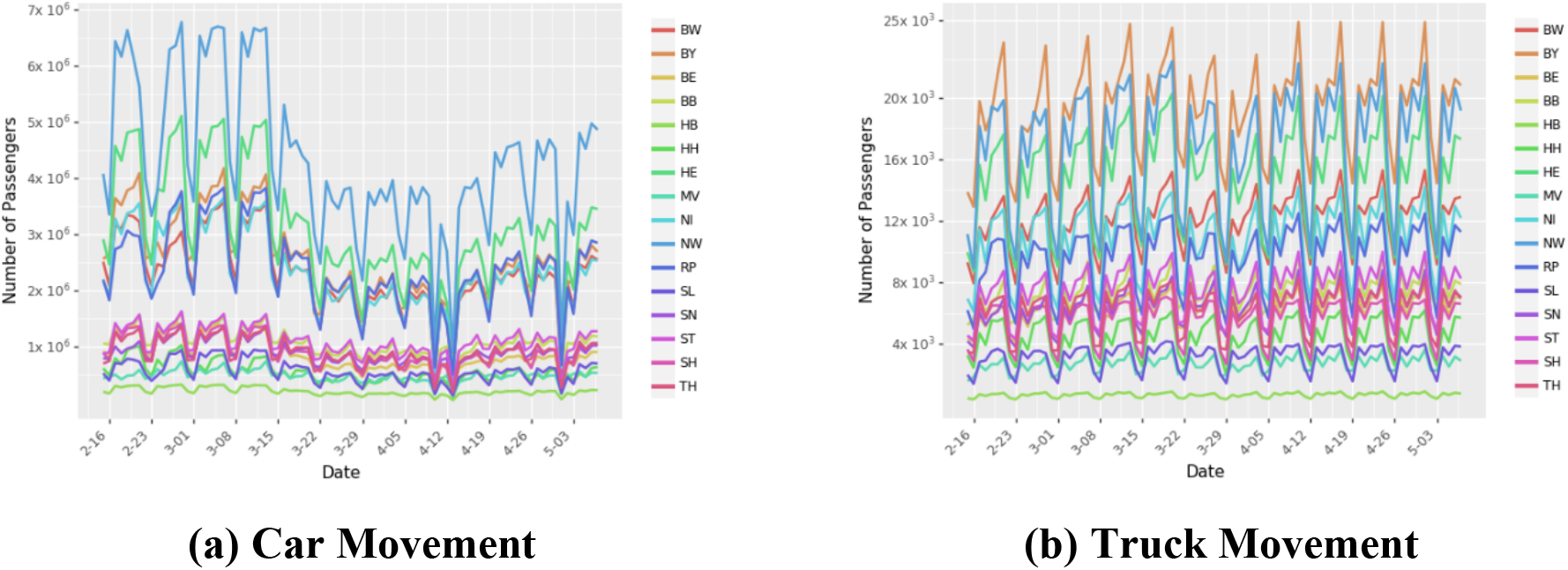
Ground transportation using cars and trucks in Germany. Panel (a) shows the number of passengers arriving at different states by car. Panel (b) shows the number of trucks arriving at different states. During the period of this study, there was no restriction on truck movement.

We used Deutsche Bahn’s timetables (http://www.bahn.com) to estimate the number of daily rail travelers moving across states in Germany and arriving from neighboring countries. To account for the changes in movement due to COVID-19 and cancelations of several trains, we adjust the number of passengers moving across states by using the community mobility data for transit stations (Figure 6).

**Figure 6.**
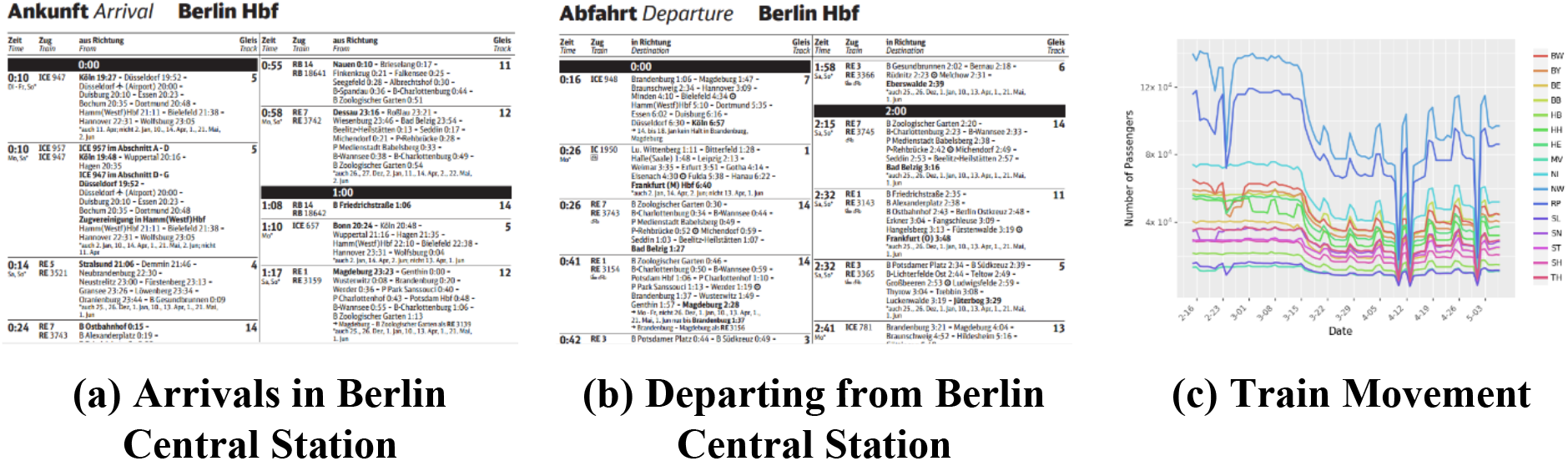
Ground transportation using trains in Germany. Panel (a) and Panel (b) show a part of the train schedules available in all the train stations. We parse these files to obtain the train schedules. Panel (a) lists the arrival time of all the trains to Berlin with departure time of all the proceeding stations for that train. Panel (b) lists the departure time for all the trains from Berlin with arrival time of all the succeeding stations for that train. We combine information from Panel (a) and Panel (b) to construct the complete route of a train. Panel (c) shows the number of passengers arriving to different states through train.

We obtained the search history of a large European bus and train comparison platform to estimate the number of passengers moving across cities (states) in Germany and passengers traveling to Germany from neighboring countries. We set bus transport to zero after March 16, 2020 as all bus movement in Germany stopped on that day.

Last, we obtained flight transportation information from the Opensky Network (20). This database utilizes Automatic Dependent Surveillance Broadcast (ADS-B) flight trajectories to identify the departure and arrival airport of a flight (Figure 7).

**Figure 7.**
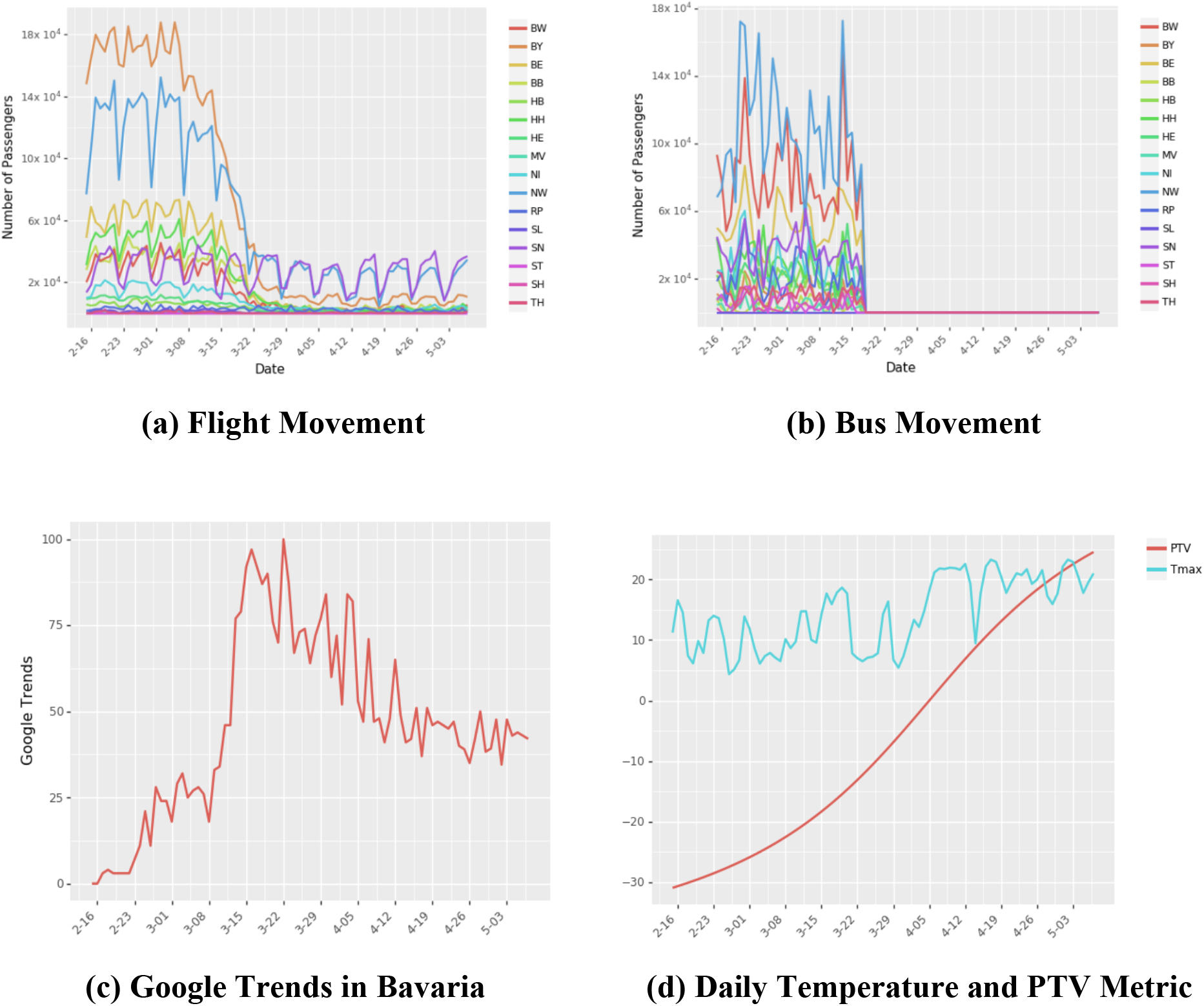
Transportation data in Germany. Panel (a) shows the number of air passengers arriving in different states in Germany. We assume 200 passengers for domestic and 500 passengers for international flights. Panel (b) shows the number of bus passengers arriving to different states from different states in Germany and neighboring countries. We assume 20 passengers per trip. Panel (c) shows the Google Trends for the search term “COVID-19 in Deutschland” for Bavaria. Google Trends numbers indicate search interest of a topic over time as a proportion of all other searches at the same time. We show the Google Trends for other states in Supplementary Material. We use cumulative Google Trends to account for increased awareness over time. Panel (d) shows the maximum temperature recorded in a day and PTV metric.

Finally, we use additional controls to isolate the effect of individual policies. We use cumulative Google Trends data for the search term “COVID-19 in Deutschland” to control for increased awareness over time (Figure 7(c)). Severe movement restrictions due to increased enforcement of these policy restrictions led to a greater sense of unease and dissatisfaction amongst some sections of the population (e.g. (21)). While such protests are small, prolonged enforcement of restrictions could increase dissatisfaction. We use weather data (max temperature in degree Celsius) from wetterkontor.de as a control to account for the propensity of the population to leave their home as summer peaks. We also include an index that measures the Propensity to Violate (PTV) NPIs using the arctan function. As the policy enforcement prolongs, the PTV index increases, in turn potentially increasing social mobility (Figure 7(d)).

## Results

### Quantifying Policy Contributions

To determine the impact of different state policies, we use a penalized linear regression (Lasso regression) model to predict changes in community mobility, *sd_i_* due to a policy *p* that is active in state *i* on a given day. We explain the regression model in detail in the Supplementary Material.

Figure 8 (a) shows the parameter estimates of various policy restrictions and controls on social mobility. We use Bootstrapping to estimate Inter-Quartile Range (IQR) for the parameter estimates (details in Supplementary Material). Figure 8 (b) shows the predictions of mobility in a sample state (Bavaria) from our model. Based on these results, we note the policies that significantly affect changes in mobility to project the number of new cases when the policy is relaxed. From the graph, we see that educational facilities closures have a very large negative effect on mobility (29.26 percentage point drop in mobility, IQR: 29.25 to 29.95). Even though it is plausible that the closure of educational facilities precipitates this drop (for example, children staying home without caregivers forces parents to work from home), we do not include this variable when projecting the lifting of policy restrictions scenarios. This is due to the potential confounding between initial awareness of the disease, population preparedness for a shutdown, and the closure of the educational facilities. The estimates for the other policy restrictions capture the average incremental effect of these policies across the 16 states in reducing mobility. The other policies listed in order of impact on mobility (estimate of percentage point drop in mobility from the regression and IQR in parenthesis) are: Contact Restrictions (12.42; IQR:12.39 to 13.32), Initial Business Closures (8.04; IQR: 8.03 to 8.65), Stay-at-Home Order (2.85; IQR: 2.88 to 3.52), Retail Store Closures (2.78, IQR: 2.78 to 3.05) and Non-essential Services Closures (2.66; 2.66 to 2.93). The model also finds that the longer the policies remain in place and restrict movement, the likelier it is for PTV to grow, which can lead to individuals breaking the policy restrictions on their own.

**Figure 8.**
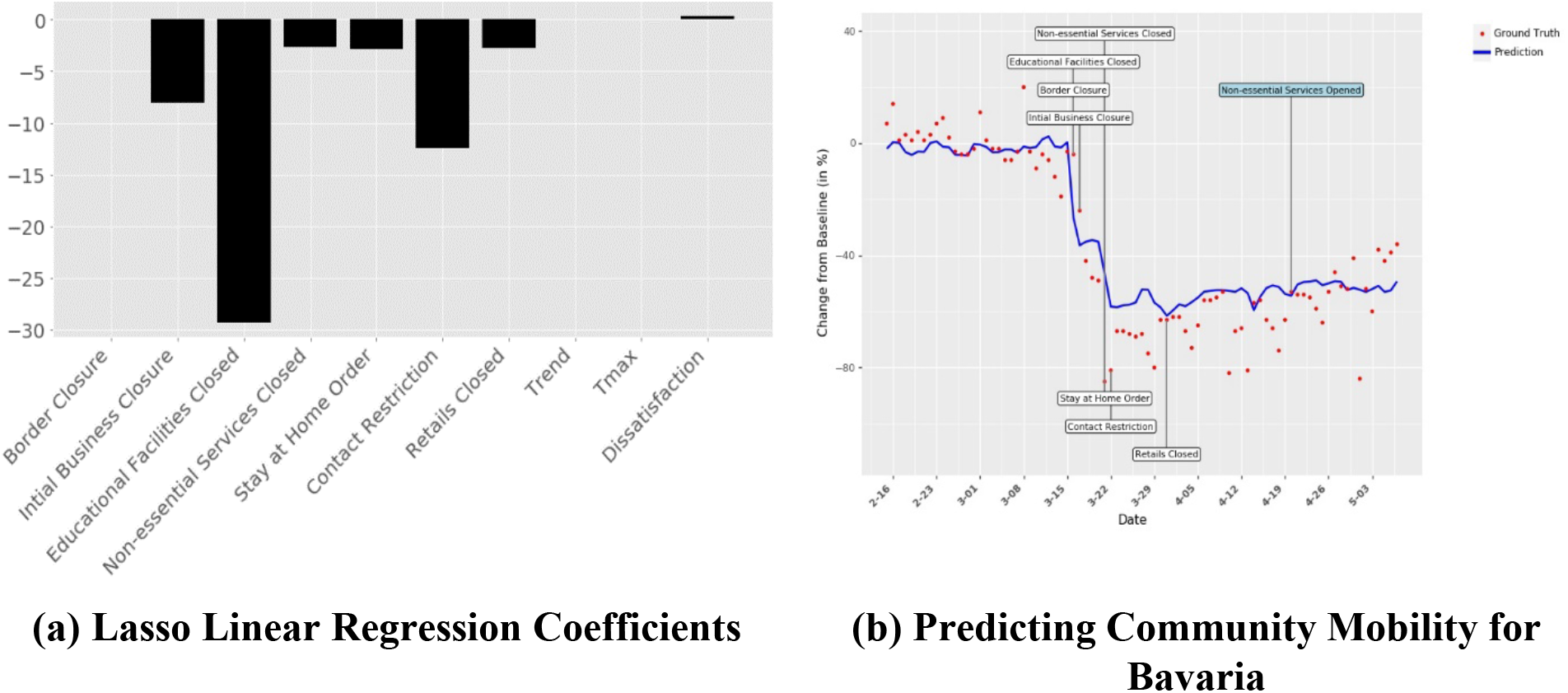
Linear (Lasso) regression model for estimating the marginal contribution of different NPIs on community mobility. Panel (a) shows the linear lasso regression coefficients for different policies. Our regression model has an R-square value of 0.72. Panel (b) shows implementation dates for different policies, true community mobility, and predicted community mobility for Bavaria. The plots for all other states are in the Supplementary Material.

### Predicting Disease Spread

We use the *predicted* mobility from the linear regression to determine the impact of social distancing on new case counts. We investigate the contribution of each policy to the mitigation of disease spread by determining the role of social distancing in the estimation of the number of susceptible and exposed individuals in a given population. We modify the SEIR model (Equations 1 – 5 in the Supplementary Material) used in (9) to include different transportation networks and predicted mobility for each state. Using the estimation procedure in (9), we find the model parameters that we use to predict disease spread for all 16 states and for Germany. We note that the model accounts for documented as well as undocumented infected cases. As shown in Supplementary Material Figure S14, the proportion of documented infected (*I^d^*) as a function of total cases increases over time. This finding comports with expectations because of the rapid increase in testing across Germany (22).

Figure 9 (a) shows the actual disease progression in Germany, the disease spread as predicted by our model in the presence of predicted social distancing, as well as disease spread as predicted by our model when mobility remained unchanged with no social distancing measures. Similar predictions for the states are provided in Figure 9 (c). We use the time period of Feb 18, 2020 – Apr 20, 2020 to infer model parameters. This period includes early stages of the *COVID-19* epidemic in Germany and the time that state policies are enacted. We use these parameters to estimate the number of daily documented cases during the time interval of Feb 18, 2020 – May 7, 2020, which corresponds to 17 days out of sample forecasts. The model finds 179,487 (IQR: 144,590 – 211,771) cumulative documented cases in Germany as of May 7, 2020 (actual reported cases: 165,991) with the estimated average error rate of 8%. Figure 9 (b) shows the amount of expected increase in the number of cases across the states of Germany and the nation, if no social distancing was observed. Across Germany one would expect a 37.8-fold (IQR: 27 to 52) increase in the number of cases without any social distancing (i.e., *Sd_i_* = 0), the effect varying significantly by states from a low of 13.7-fold (IQR: 7 to 21) in Berlin to a high of 45.2-fold (IQR: 31 to 63) in Bavaria.

**Figure 9.**
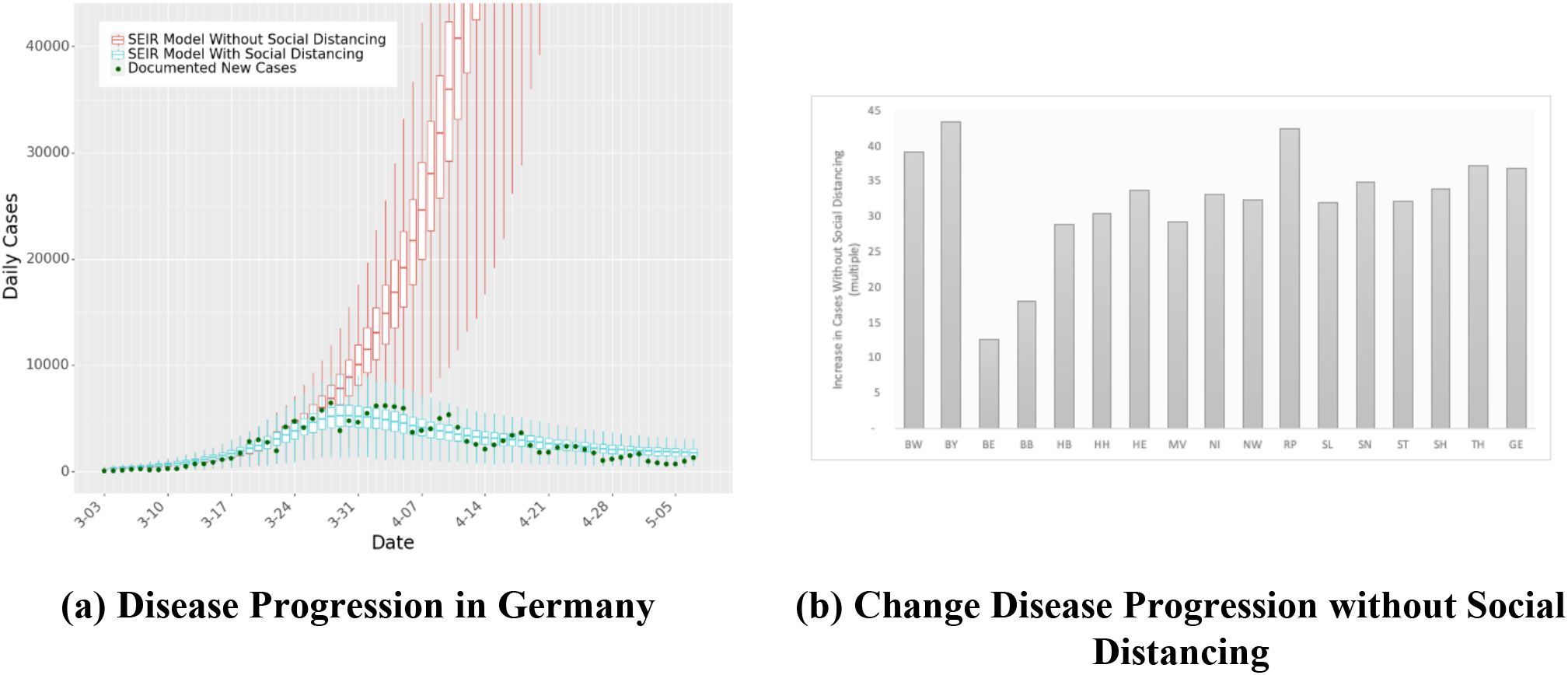
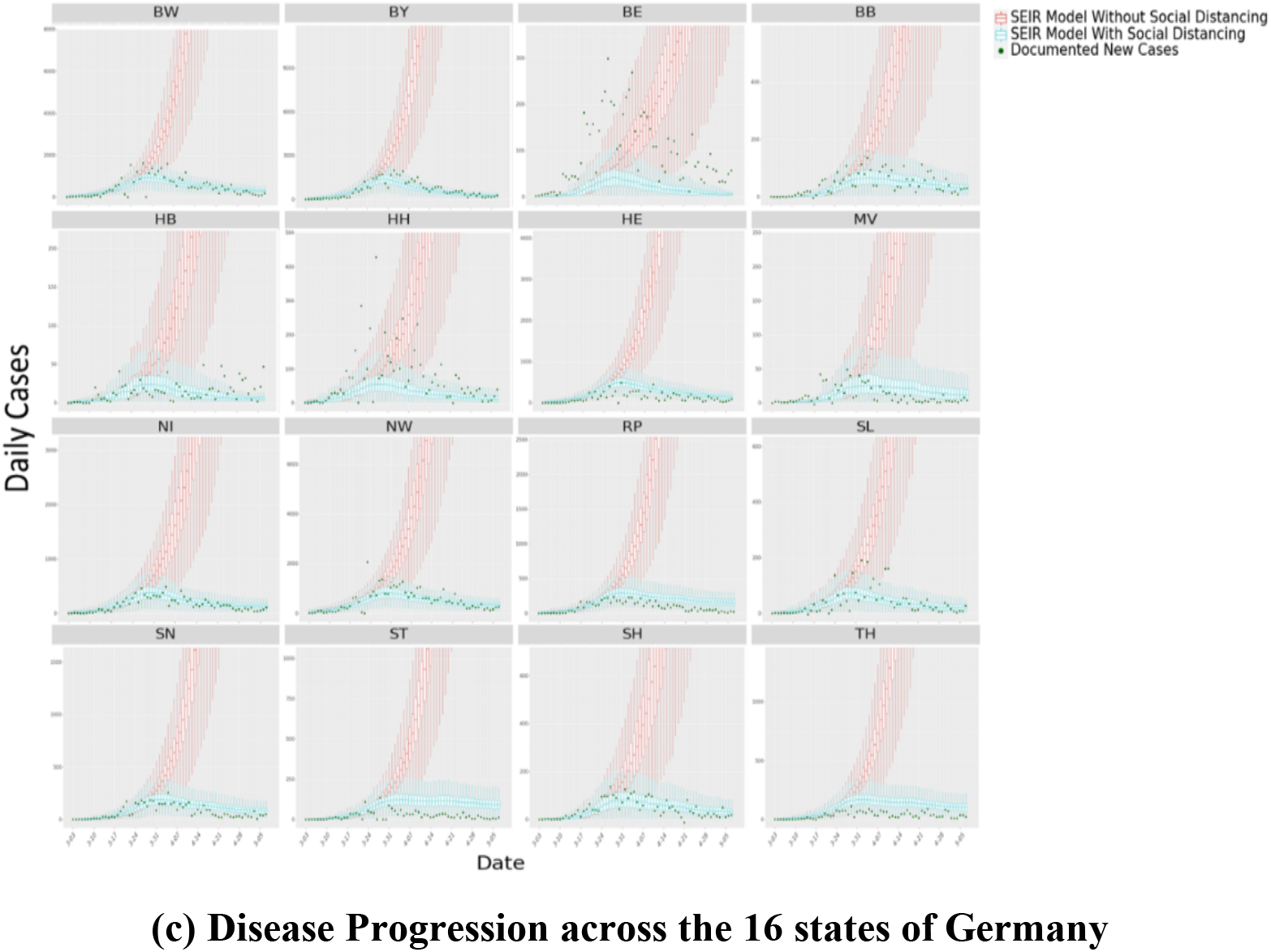
SEIR Model Predictions. Panel (a) shows the predictions of new case counts in Germany with and without any distancing measures, against actual cases for the same time period. Panel (b) charts the expected increase in cases (multiple) without any distancing measures. Panel (c) plots the predictions of new case counts with and without any distancing measures, against actual case numbers for all states. Our predictive accuracy is higher for states with high case numbers such as Baden-Württemberg (BW) and Bavaria (BY) than for the small city-states Berlin (BE), Bremen (HB), and Hamburg (HB). We note that the last 17 days of Panels (a) and (c) correspond to out-of-sample forecasts from the SEIR model.

### The Lifting of Restrictions

We simulate what-if scenarios to determine the impact of lifting restriction on new cases in each state. Under the scenario that a restriction has been relaxed while others remained operational, we forecast mobility using Equation (11) in the Supplementary Material. We subsequently project the new case count using the predicted mobility. This step is repeated across all restrictions, each relaxed individually. Due to the confounding noted earlier, we do not report the case of educational facilities being reopened.

In *Scenario 1*, we project all changes from April 21, 2020 to June 2, 2020, assuming that the restriction was relaxed on *April 21, 2020*. Figures 10 (a) and (c) shows the projections of case counts over a six-week period if a restriction was relaxed exclusively. Figures 10 (b) and (d) show *Scenario 2*, which projects all changes from April 21, 2020 to June 2, 2020, assuming that the restriction was relaxed on *April 28, 2020*. Because the two scenarios are exactly one week apart, it allows us to determine the impact of delaying the lifting of a restriction by one week. From the analysis, the lifting of contact restrictions, i.e., the rule limiting movement in public spaces, had the biggest impact on new case counts. Compared to keeping the restrictions in place, lifting contact restrictions will result in a 51.5% (IQR: 51.0–52.1%) increase in daily case numbers in scenario 1 and a 27.4% (IQR: 26.8–27.8%) increase in scenario 2. However, lifting restrictions on initial business closures leads to a 29.2% (IQR: 28.6–29.7%) increase in daily case numbers in scenario 1 and a 15.8% (IQR: 15.4–16.3%) increase in scenario 2. Easing nonessential service closures increases daily case numbers by 6.6% (IQR: 6.2–7.0%) in scenario 1 and 3.7% (IQR: 3.2–4.0%) in scenario 2, and the opening of retail outlets increases daily case numbers by 5.6% (IQR: 5.1–6.0%) in scenario 1 and 3.4% (IQR: 3.0–3.8%) in scenario 2. These results show that NPIs have differential impacts on lowering disease spread, and suggest a measured approach to lifting restrictions. For example, the opening of retail outlets could be balanced by maintaining the restrictions around limiting the number of individuals in a given place or store (e.g., controlling entry) – thereby allowing for the resumption of economic activity while limiting the risk of contagion.

Figure 10 (e) shows the increases in the expected number of cases if a restriction was lifted on April 21 versus on April 28. Delaying the lifting of certain restrictions by one week could also have a significant impact on the total case counts. This occurs not only due to the delay, but also because the number of infected individuals that a person could come in contact with decreases over the week. For example, delaying the lifting of contact restrictions by one week reduces the number of new cases over the six-week forecast by on average 19% (IQR: 16% to 21%). We also observe that lifting restrictions on the opening of retail outlets and non-essential services leads to an average 2% (IQR: 0% to 6%) and 3% (IQR: 0% to 4%) increase in total case numbers over the six-week forecast period.

**Figure 10.**
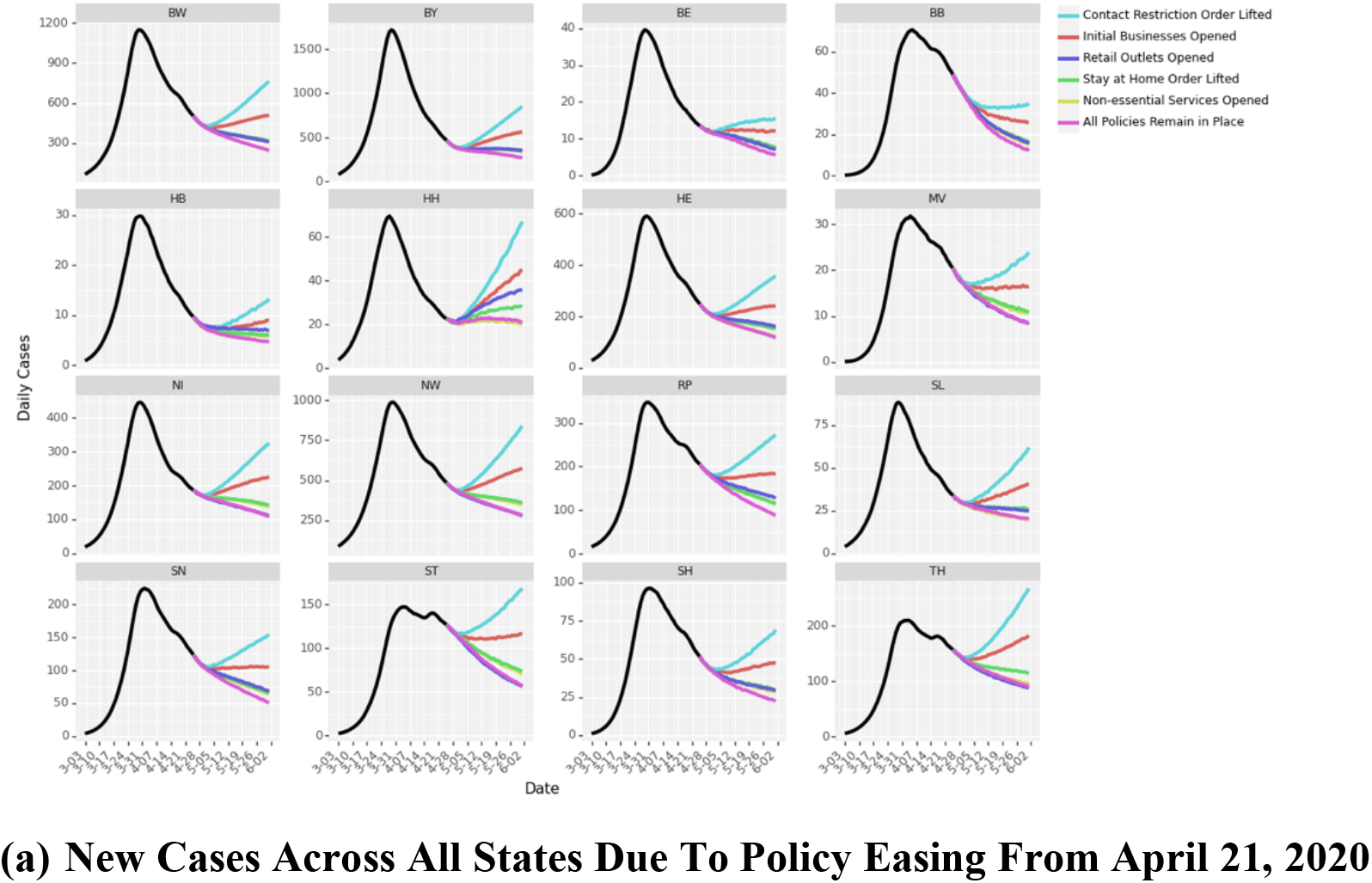
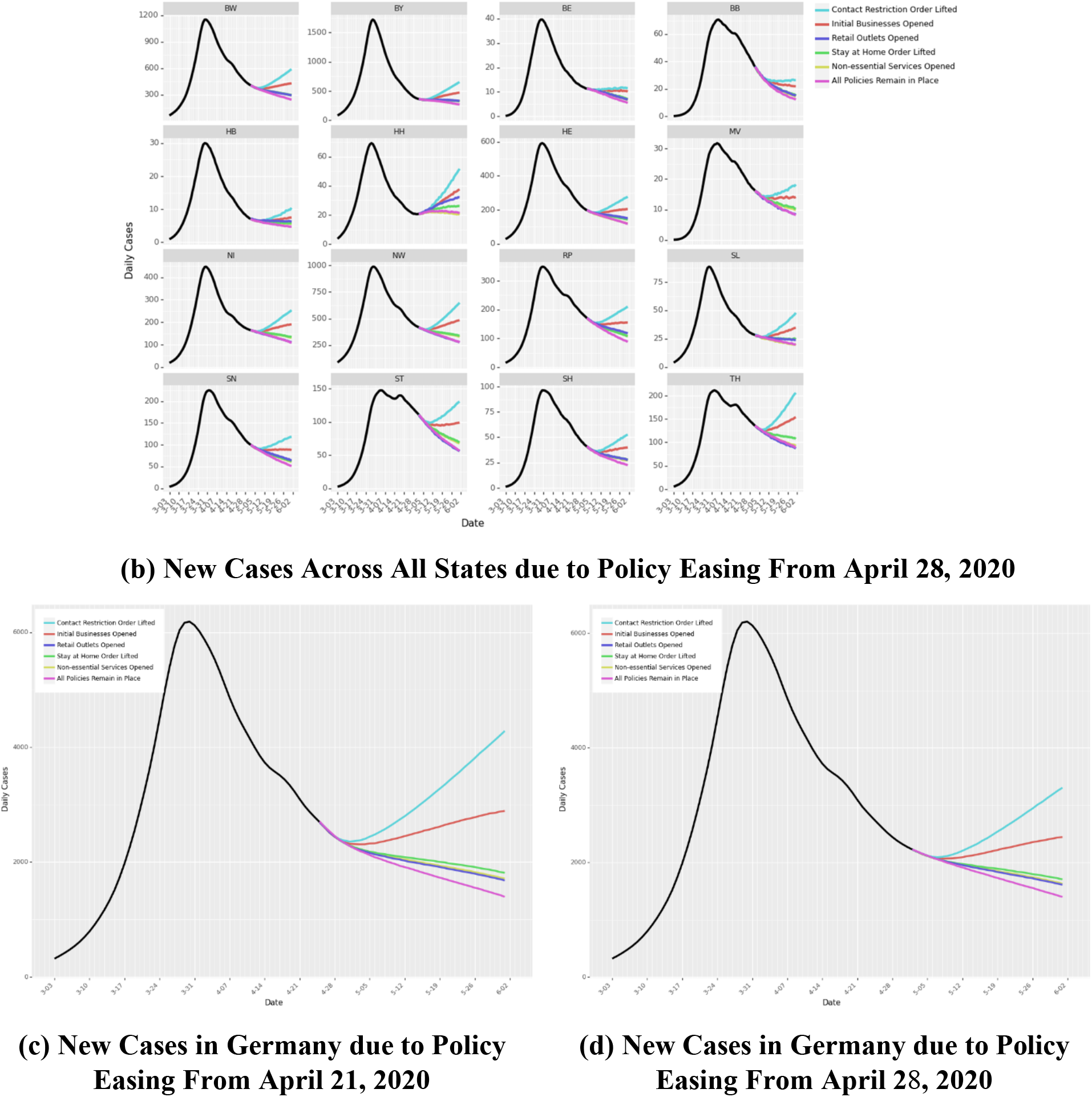
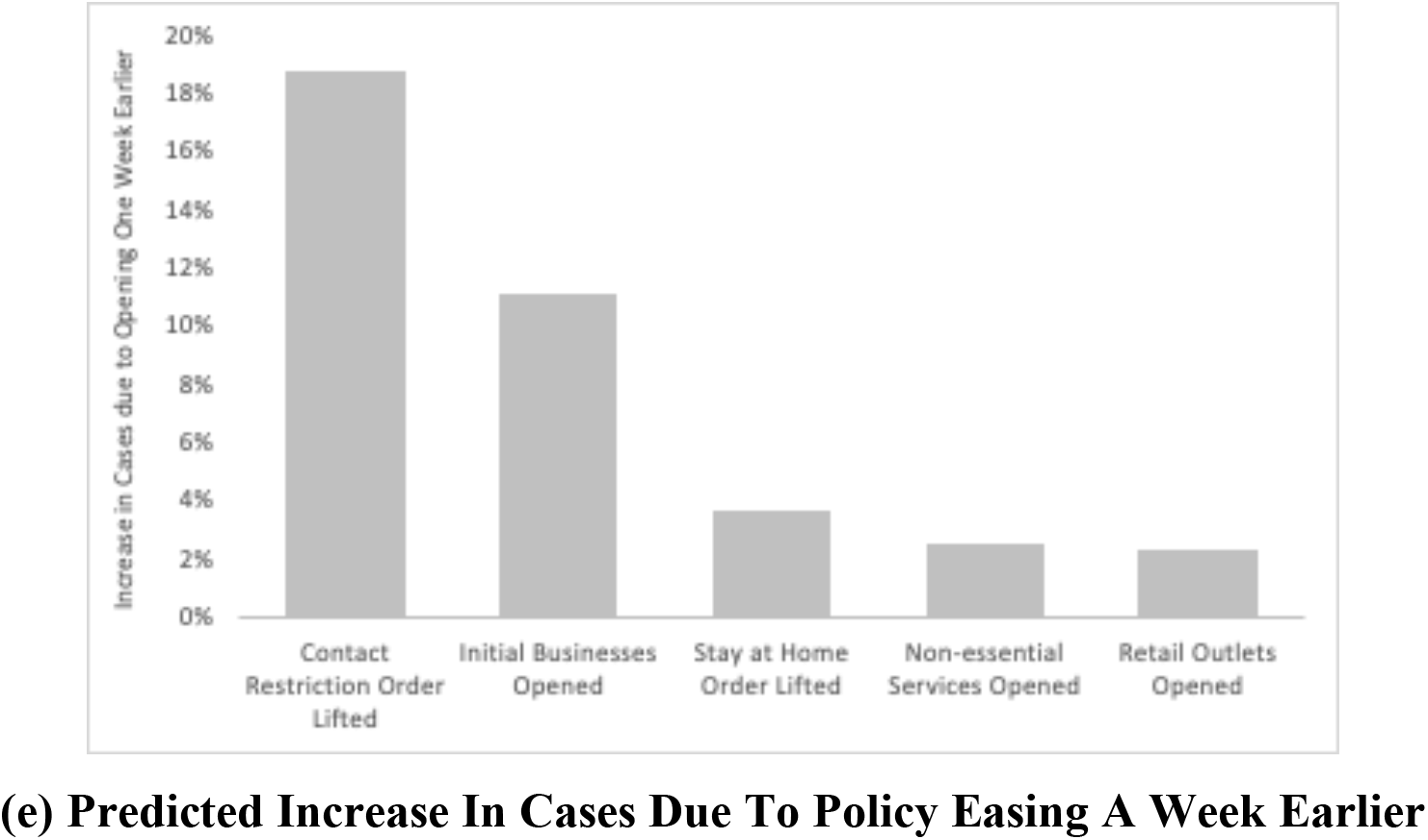
Lifting of Policy Restrictions. Panel (a) and (b) show the predictions of new case counts across all states of Germany due to lifting of policy restrictions from April 21, 2020 to June 2, 2020. In Panel (a) policy restrictions are relaxed, one-at-a-time on April 21, 2020. In Panel (b) policy restrictions are relaxed, one-at-a-time on April 28, 2020. Panel (c) and (d) illustrate the same for all of Germany. Panel (e) charts the predicted increase in cases due to policy easing a week earlier.

## Discussion

This study explores the role of NPIs in reducing the spread of COVID-19. We extend the spatiotemporal SEIR model in (9) by incorporating daily social distancing numbers from transportation data and mobility patterns. Our model finds that without NPIs in place, COVID-19 cases would likely have shown a 37.8-fold increase across Germany. We also investigate the marginal impact of each of the various NPIs implemented by state governments in Germany by determining the differential impacts of the policies on reducing social mobility. We relate these reductions to disease spread, reconstructing patterns of spread across the 16 states. Finally, we forecast new cases when policies are relaxed, one-at-a-time. We find that certain policies have a larger impact on disease spread than others. Our model forecasts find that early relaxation of some NPIs could lead to an increase in the number of cases, potentially leading to a second wave. This observation is confirmed by an estimated increase in the effective reproduction number *R_e_* (Supplementary Material). We also compare case counts if the policies were relaxed with a one-week delay. Keeping some NPIs in place for an extra week could reduce total COVID-19 cases by up to 19% (as of June 2, 2020). The results confirm that policy restrictions are not all equal in their ability to affect disease spread. The policy of restricting mass gatherings (Contact Restriction) is estimated to be the most effective NPI to contain COVID-19, followed by closures of various businesses and stay-at-home orders. Due to this variation in effect, it is advisable to lift restrictions with minimal effects first, gradually easing restrictions that potentially lead to higher case numbers. This study presents a comprehensive quantitative analysis that includes individual effects of NPIs on the transmission of COVID-19. To the best of our knowledge, this is the first study that uses variations in policy interventions by governments to discover their differential impacts at reducing mobility that in turn reduces disease spread. Prolonged lockdowns and restrictive policies can have devastating social and economic consequences. However, opening too soon could result in rapid disease spread. Therefore, governments need to develop cautious approaches to lifting restrictions in a bid to return to normalcy (23). The approach presented in this paper allows for a deeper understanding of the policy effects on mitigating the spread of COVID-19. The forecasts of disease spread when NPIs are (partially) loosened guide policymakers towards the appropriate strategy when reversing the interventions.

## Data Availability

Data available upon request

## Notes

### Competing Interest Statement

The authors have declared no competing interest.

### Funding Statement

No funding

### Author Declarations

No IRB required

